# Sex-stratified analysis of factors associated with attrition intent from the General Internal Medicine physician workforce in Switzerland

**DOI:** 10.1101/2025.09.04.25335096

**Authors:** Tabea Leitner, Isaac Egger, Sven Streit, Jeanne Moor

## Abstract

**Background:** The healthcare sector has a shortage of physicians. Strategies to better retain medical professionals in the workforce in General Internal Medicine must rely on an in-depth understanding of factors associated with wanting to quit their job. Here, we investigated sex-specific associations of workplace-related and personal factors associated with wanting to quit work among physicians.

**Methods:** In a cross-sectional questionnaire among physicians working in General Internal Medicine in Switzerland, we assessed personal and workplace-related factors in association with the desire to quit their job. The outcome variable of wanting to quit one’s job was dichotomized from a 6-point Likert scale. We performed sex-stratified analyses by Wilcoxon rank sum test, Chi-square test and multiple logistic regression adjusting for demographic variables.

**Results:** This study included 682 physicians, 278 (41%) men and 404 (59%) women aged 37±11 years (mean ± standard deviation). A majority of 78% men and 75% worked in hospitals. Overall, a desire to quit their job was prevalent in 33% of respondents of either sex. Almost all workplace-related items were associated with the probability of wanting to quit among both sexes: Having a good network, mentoring or supervisor’s support were associated with a lower probability of wanting to quit, whereas having a bad work-life balance or dissatisfaction with autonomy at work. Problematic or workplace inclusiveness and experienced gender-related discrimination at work were associated with a higher probability of wanting to quit in univariable analysis in both sexes. The main sex difference was that the associations of workplace inclusiveness or gender discrimination with wanting to quit were robust to adjustment by type of workplace and language region in men, whereas in women upon multivariable adjustments the associations disappeared. Finally, in men (but not women) having no adequate childcare was more likely to desire quitting their job.

**Conclusion:** This study identified several factors of which some may exert a causal relationship with the desire to quit a physician job. Modifying such factors by interventions may ultimately increase the likelihood of a physician continuing working in his profession.

## Introduction

The World Health Organization (WHO) reports a healthcare workforce deficit in many European countries, including in Switzerland, which is particularly evident in general internal medicine (GIM).(1,2) This workforce deficit is only in part explained by the demographic development of the ageing general population, their increasing healthcare participation, an increasing share of physicians working part-time, and many general practitioners about to reach retirement age.(3,4) In fact, physicians are also at risk of attrition from the workforce. Studies among physicians worldwide showed that 11-55% of physicians intend to leave the profession.(5–7) Similarly, in Switzerland one third (34%) of Swiss physicians reportedly consider at least sometimes to leave the workforce early.(8)

Known drivers of physicians’ attrition risk include on the one hand personal factors such as parenthood or a pathological mental well-being.(9–11) On the other hand, physician attrition is driven by workplace-related factors such as long working hours, work-life conflicts, having no professional network, a lack of mentoring or supervisor’s support, gender discrimination and an insufficient workplace inclusiveness.(4,9–15) A striking gap in knowledge remains, however, how disproportionally these drivers of attrition affect male and female physicians. For instance, parenthood is considered a key factor for the risk of attrition from the workforce among female physicians but not in male physicians.(10,14) Indeed, parenthood is a highly gender-sensitive obstacle for professional advancement, affecting female physicians more than male physicians due to lack of support by the employer, absence of adequate childcare and maternal discrimination. (14) Designing future strategies to retain physicians in the active workforce requires a better understanding of potentially modifiable mechanisms that drive physician attrition from the workforce. The WHO reported that sex-disaggregated data regarding how to retain medical workforce is lacking, which includes gender-specific measures in working conditions, gender differences in terms of burnout and attrition and family-friendly measures like childcare support or part-time(16).

In the present study, we analyzed a dataset of a survey among Swiss physicians in GIM, with a focus on the risk of physician attrition from their job. We performed in-depth analyses to determine sex-specific associations of workplace-related and personal factors with attrition risk to better understand the potential key drivers of attrition in the GIM physician workforce, and to obtain an overview of the underlying sex and gender differences.

## Methods

### Design and population

We previously reported on a cross-sectional, web-based survey among GIM physicians in Switzerland (17–19). In brief, participants were recruited via 14 hospitals and six ambulatory or primary care institutes in Switzerland, via a newsletter of the Swiss Society of General Internal Medicine, an unpaid advertisement in the Swiss journal *Primary Hospital Care* or an invitation e-mail sent to members of the Swiss Young General Practitioners Association (JHaS). The study was waived by the ethics committee of the Canton of Bern (identifier: Req-2021-01085). Participants were informed about the purpose of the study and provided informed consent for voluntary and anonymous participation.

### Data collection

The survey was hosted on the website www.Surveymonkey.com for participation between December 2021 and April 2022. Data collection strategy was previously described in detail (17).

Outcome of the present secondary analysis was the desire for quitting the job as an estimate of physician attrition risk, assessed using a 6-point Likert scale: “I want to quit my work…”, with options ranging from 1 (absolutely not) to 6 (absolutely). For the analysis, the responses were dichotomized (“Yes” vs. “No”).

As exposure variables or potential confounders, we included additional data in the analysis including respondents’ demographic information, personal and family information, and their professional situation. Workplace inclusiveness was evaluated by 3 questions from the 19-items questionnaire on organizational culture by Hall et. al. (22). Gender discrimination was assessed using 6 questions developed earlier(17).

The survey was offered to participants in German and French language, after pilot-testing and back-and-forth translation.

### Statistical analyses

Variables are reported as mean (standard deviation) or median (interquartile range) for continuous data, and n (%) for categorical data. Student’s t-tests, Wilcoxon rank-sum test were used for continuous variables as appropriate, and Chi-square test for categorical data. Logistic regression models were used to model the outcome of physician attrition risk. Exposure variables were included in both univariable and multivariable models. Separate multivariable logistic regression models were calculated for each exposure of interest. A priori covariates were age, type of workplace and Swiss language region. Sex was used as stratification variable. No power analysis was performed for the present post-hoc analysis. Analyses were conducted with RStudio v. 2023.06. P-values <0.05 were considered as statistically significant. We made no adjustments for multiple testing.

## Results

### Demographics

In total, 684 physicians completed the survey. Two respondents (0.3%) with non-binary gender identity with specified gender ‘other’ were excluded from all sex-stratified analyses to preserve anonymity. Participant characteristics are summarized in Table 1. Among the participants, 404 (59%) were women and 278 (41%) were men. Mean age among women was 36 (SD 10) years and among men 39 (SD 12) years. A majority of participants (76%) worked in hospitals, while 21% worked in private practice.

**Table 1.** Respondent characteristics.

|  | **N** | **Men,**<br>**N=278** | **Women,**<br>**N=404** |
| --- | --- | --- | --- |
| **Age in years** | 682 | 39 (12) | 36 (10) |
| **Swiss language region** | 682 |  |  |
| German-speaking |  | 197 (71%) | 288 (71%) |
| French-speaking |  | 73 (26%) | 111 (27%) |
| Italian-speaking |  | 8 (2.9%) | 5 (1.2%) |
| **Type of workplace** | 682 |  |  |
| University hospital |  | 104 (37%) | 155 (38%) |
| Other hospital |  | 115 (41%) | 152 (38%) |
| Private practice with $\geq 3$ colleagues | | 23 (8.3%) | 43 (11%) |
| Private practice with $< 3$ colleagues | | 29 (10%) | 36 (8.9%) |
| Other place |  | 7 (2.5%) | 18 (4.5%) |
| Data are reported as n (%) or mean (standard deviation) |  |  |  |

### No sex differences in risk of physician attrition

Among physicians of each sex, 33% of participants answered the survey with a positive response when questioned if they want to quit their job (Table 2, sex difference p=0.945). Missing data were present in 63 men (23%) and 68 women (17%).

**Table 2.** Sex-stratified desire to quit the job.

| **Characteristic** | **N** | **Men,**<br>**N=278** | **Women,**<br>**N=404** | **p-value** |
| --- | --- | --- | --- | --- |
| Desire to quit | 551 | 71 (33%) | 110 (33%) | 0.945 |
| Unknown |  | 63 | 68 |  |

Comparison by Chi-squared test

### Limited associations between intrinsic personal characteristics and risk of physician attrition

Among female physicians, there was no association between any personal factor and wanting to quit their job in univariable and adjusted analyses of several variables, including perceived spouse’s support for one’s career, being a parent, or own age at the birth of the first child (Figure 1, upper panel). Among male physicians, reporting to have no adequate childcare was associated with wanting to quit one’s job in both univariate analyses (OR: 3.15, 95% CI 1.22-8.20, p=0.018) and after adjustment for confounders in multivariable analysis (OR: 3.64, 95% CI 1.24-11.2, p=0.020) (Figure 2, upper panel). In addition, with an increasing number of children among male respondents, the likelihood of wanting to quit the job decreased in the univariate analysis (effect per child: OR 0.75, 95% CI 0.59-0.95, p=0.019). However, this association was not robust to adjustment for confounding (multivariable analysis, effect per child: OR 0.96, 95% CI 0.70-1.30, p=0.79). Other variables such as spouses’ support or being a parent were also not associated with wanting to quit the job among male physicians, similarly as in female physicians.

**Figure 1.**
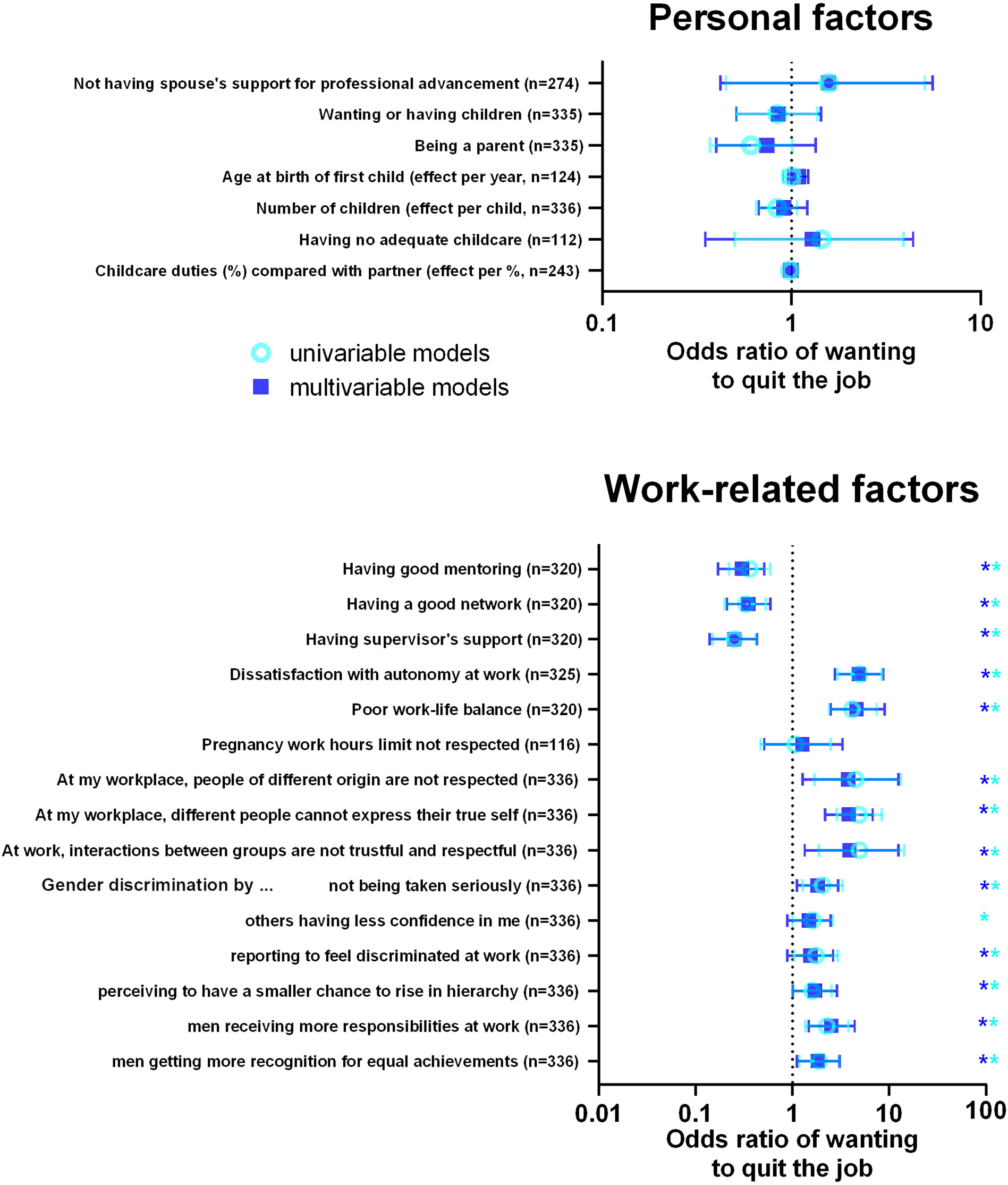
Factors associated with wanting to quit the job among female physicians. Forrest plot of the odds ratios of wanting to quit the job among female physicians. Each line represents an univariable model (light blue) or multivariable logistic regression model (dark blue) adjusted for age, Swiss language region and type of workplace. Stars indicate significant associations (p<0.05).

**Figure 2.**
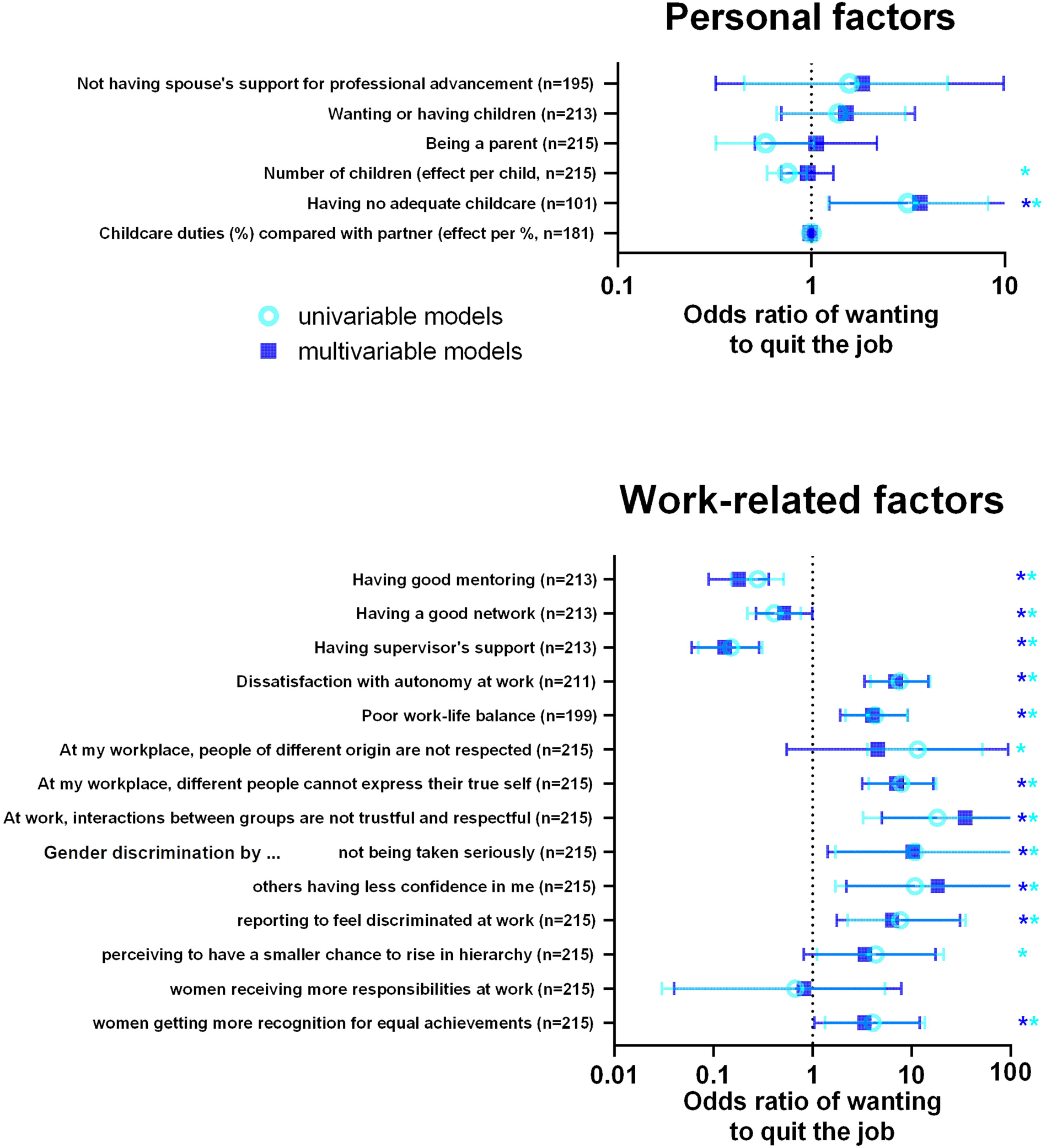
Factors associated with wanting to quit the job among male physicians. Forrest plot of the odds ratios of wanting to quit the job among male physicians. Each line represents an univariable model (light blue) or multivariable logistic regression model (dark blue) adjusted for age, Swiss language region and type of workplace. Stars indicate significant associations (p<0.05).

### Work-related factors are associated with a risk of physician attrition among both sexes

In contrast to the limited associations between personal factors and wanting to quit the job, several work-related factors were associated with wanting to quit their job among physicians of both sexes in the univariable and the multivariable models (Figures 1-2): Dissatisfaction with autonomy at work was associated with a higher likelihood of wanting to quit the job among respondents of both sexes (OR: 4.85, 95% CI 2.76-8.67 in women, OR: 6.87, 95% CI 3.33-14.8 in men). Similarly, having a poor work-life balance were both associated with a higher likelihood of wanting to quit the job among respondents of both sexes (OR: 4.6, 95% CI 2.49-8.93 in women, OR: 4.03, 95% CI 1.91-9.17 in men). By contrast, having a good network, good mentoring, or having supervisor’s support were all associated with a lower likelihood of wanting to quit the job among respondents of both sexes.

Regarding workplace inclusiveness, three items were associated with wanting to quit the job among physicians of both sexes. These items were observing disrespect towards people of different origin, different people’s inability to express their true self, and interactions between different groups being not trustful and respectful. Most of these associations remained robust to multivariable adjustment, with the exception that the association between disrespect towards people of different origin and wanting to quit remained robust to adjustment in women (OR: 3.76, 95% CI: 1.27-12.5, p=0.021) but not in men (OR: 4.56, 95% CI: 0.55-94.8, p=0.2). Overall, these findings demonstrate a significant connection between impaired workplace inclusiveness and a desire of physicians among both sexes to quit their job.

### Association between gender discrimination at the workplace and wanting to quit the job

Among participating women, all items assessing gender discrimination at the workplace were associated with wanting to quit the job in the univariable analysis, and most also in the multivariable analysis (Figure 1, lower panel). Among men, the majority gender discrimination items also showed an association with wanting to quit the job in univariable and multivariable analyses (Figure 2, lower panel). An intriguing finding was that among female physicians, reporting that male physicians more responsibilities at work was associated with wanting to quit the job (OR: 1.84, 95% CI: 1.11-3.07, p=0.018). Among male physicians, there was also evidence that perceiving female physicians to receive more responsibilities was associated with wanting to quit the job (OR: 3.35, 95% CI: 1.04-12.0, p=0.048). Overall, these results demonstrate that physicians of both sexes showed largely congruent associations between gender-related discrimination and their desire to quit their job.

## Discussion

### Principal findings

In this large national cross-sectional study, we aimed to gain evidence of potential mechanisms underlying physician attrition in GIM, with an aim of identifying challenges experienced by physicians of either sex. Overall, we found that 1 in 3 GIM physicians in Switzerland had stated a desire to quit their job, highlighting the pressing nature of the topic of workplace attrition in physicians Switzerland. In sex-stratified analysis of potentially associated personal factors, we found that an overwhelming majority of factors related to workplace environment such as workplace inclusiveness, impaired work-life balance, dissatisfaction with autonomy on the job and gender-related discrimination were associated with the desire to quit the job among physicians of both sexes. Notably, almost no intrinsic personal factors were associated with the desire to quit, except that having inadequate childcare were associated with a desire to quit among male physicians.

### Comparison of the findings with the literature

The main findings of this study are supported by previous international and national literature showing a prevalence of the intention to leave the medical field in around 20%.(7,10,20) The fact that our findings are higher than the average findings in literature could be a result of differently formulated questions: In our questionnaire, we asked participants about their *desire* to quit their job, whereas other studies used stronger wording and asked about *intention* to leave. The intention to leave the medical field has been found to correlate with actual departures.(6)

### Work-life conflicts

Our data showed that poor work-life balance was one of the factors associated with a desire to quit one’s job as a physician. This is in line with data from the VSAO survey: Compatibility of work with childcare was the second most important reason to leave the workforce, after the number of working hours. Further associated factors were being dissatisfied with one’s autonomy at work. In addition, our previous analysis revealed that gender-specific work-life conflicts can have a substantial impact on physician wellbeing.(19)

### Personal Factors and risk of physician attrition

In our study, relationship and family related factors did not show an association with wanting to quit the job among female physicians, which distinguished them from male physicians. Not having adequate childcare services was linked to wanting to quit the job among men but not among women, which is intriguing when looking at literature: In Switzerland, physicians tend to take on traditional gender roles when having children: While most physician-fathers continue full-time after birth, physician-mothers cut back to part-time work and assume more family duties, since women are still primarily responsible for childcare and household.(21) Also, most female physicians’ spouses work full-time, whereas a much smaller share of male physicians’ spouses do.(21,22) Therefore, one would assume that adequate childcare might be more important for female physicians to reduce work-life conflicts. However, our findings might reflect the fact that men working part-time is still less socially accepted.(23) Furthermore, traditional gender roles are breaking down in Switzerland so that young men increasingly take on household and childcare duties.(24)

### Workplace inclusiveness

Impaired workplace inclusiveness was linked to a higher likelihood of wanting to quit in the present study. These findings are in accordance with the results of another Swiss study, which has demonstrated that workplace discrimination is significantly associated with wanting to quit the job among physicians.(13) That the more exposed physicians are to different forms of workplace discrimination, the more likely they are to consider quitting the job.(13,25) Further, with regard to regarding immigration and cultural diversity, it is crucial to diversify healthcare workforce since workplace inclusiveness prevents health-related societal inequities and improves access to health care and health outcomes in social minorities.(26,27) Therefore, recruitment strategies should be implemented that retain physicians with different personal and ethnic backgrounds.

### Gender discrimination

Gender-based discrimination was also associated with wanting to quit one’s job, regardless of participant gender. In Switzerland, 31.8% of women versus 6.8% of men reported having experiences acts of discrimination, sexism or gender inequalities.(15) Workplace-related gender discrimination affects women’s hiring, promotion, and career development′ leading to the phenomenon called the leaky pipeline.(17,25,28) Also, literature demonstrates that female physicians are being held to higher standards, receive less recognition for their work, and being negatively and less formally treated by colleagues more often than their male colleagues.(29) Although significantly higher rates of gender discrimination are reported by female physicians, male physicians can also be affected by this type of discrimination. In male physicians, negative reactions regarding parenthood and childcare needs are regarded the main factor contributing to workplace-related gender discrimination.(30) Our finding might indicate that, in contrast to female physicians who seem more “used to” or are expecting experience gender discrimination to be rather the norm, male physicians appear also to be sensitive about phenomena of gender-related discrimination.

### Strengths

Strengths of this study include the comprehensive description of both personal and work-place related factors in association with wanting to quit the job. Also, the study involved a large sample of 684 participants and participants from all regions of Switzerland. Further, this study has a good chance to distinguish between sex or gender differences, as the survey was completed by respondents of a relatively balanced gender ratio. Beyond that, the identified differences in factors associated with wanting to quit the job among female and male physicians highlight an importance for the workforce to overcome negative views on parenthood and gender-specific discrimination. Thus, the present findings provide a strong incentive for targeted measures to promote structural change in Swiss hospitals.(31)

### Limitations

The present study has some limitations. To begin with, the survey response rate cannot be ascertained since the number of viewers of the survey advertisements remains unknown and no website views were measured. However, features such as the previously reported physician well-being index in the current study population(19) were comparable to that of previous Swiss studies which had a response rate of 40%.(12,13) This suggests that, for example, physicians experiencing poor mental health were not disproportionately participating in the present study. Additionally, causality could not be determined solely based on the reported associations, as is the case with all observational studies.

### Outlook and Implications

Our findings reflect a systemic problem in the Swiss medical field. One out of 3 physicians considering leaving the profession may become a threat to the health care system. According to the Swiss Health Observatory (Obsan), by 2030 30% of the needed consultations in the Swiss healthcare system can no longer be covered.(4) Furthermore, the physician shortage decreases quality of care, increases the risk for medical errors and decreases patient satisfaction.(10) Up until now, Swiss healthcare depends on an increasingly foreign workforce, since only about half of the available positions for residents in Switzerland can be filled by domestic physicians. However, it could eventually become increasingly difficult to recruit foreign personnel as Germany, France and other European countries are taking measures to minimize the “brain drain” themselves. Soon most employees will belong to Generation X and Z, requiring employers to adapt to the new generations to retain talent. Meaningful actions to preserve the workforce could include: readily available mentoring for employees experiencing discrimination, education on unconscious bias in leaders, establishment of equal career opportunities for all physicians, networking spaces, reduced weekly working hours, relief of administrative tasks, making part-time employment and job-sharing more attractive, and making childcare more affordable.(26,32)

### Conclusion

Overall, we found that one in three Swiss GIM physicians reported wanting to quit, in association with workplace conditions such as poor work-life balance, limited autonomy, gender discrimination, and lack of inclusiveness, while intrinsic personal factors played no relevant role. These findings can help to guide reforms in organizational culture to prevent physician attrition and maintain healthcare system stability.

## Acknowledgements

The authors acknowledge the contributions of the survey participants, and the support by Swiss Society of Internal Medicine, Swiss Young General Practitioners Association (JHaS), Primary Hospital Care and hospitals involved with distribution of the survey.

## Data availability

All data produced in the present study are available upon reasonable request to the authors.

